# Wastewater pathogen composition in Atlanta, GA prior to and during the 2026 FIFA World Cup

**DOI:** 10.64898/2026.11.03.26366867

**Authors:** Nina Raymo, Colleen S. Kraft, Dalia Gulick, Jessica Ingersoll, Madeleine Stoffle, Orlando Sablon, Jane Logan, Erin Haslett, Christine L. Moe, Marlene K. Wolfe, Lorenzo D. Freeman, Thomas Bourne, Vontressia Lawson, Paige E. Harton

## Abstract

Mass gatherings can have many impacts on public health, including facilitating the spread of infectious diseases. Alternative surveillance modalities, such as wastewater-based surveillance (WBS), can complement traditional clinical surveillance. We utilized WBS to examine the influence of the 2026 FIFA World Cup (FWC’26) on the range of infectious pathogens in Atlanta, GA surrounding matches held in the city. Samples (n = 102) were collected from three water reclamation facilities between April and July of 2026 (34 collection days) and tested using the BIOFIRE^®^ WATCHFIRE™ Respiratory (19 targets) and Gastrointestinal (22 targets) PCR Pathogen Panels. Primary data analysis included a temporal comparison of wastewater pathogen profiles before (April 1 to June 10; n = 60) and during (June 15 to July 15; n = 30) the FWC’26 across the three facilities using a modified Poisson model with Bonferroni correction. There were no statistically significant differences in target detection between the two periods. Samples collected after the FWC’26 (July 20 to 29; n = 12) were included in a sensitivity analysis which showed a statistically significant increase in SARS-CoV-2 (RR = 3.21, 95% CI = 1.40-7.39; p = 0.01, p-adjusted = 0.048) during and after the tournament compared to before. However, this coincided with regional and national increases in SARS-CoV-2 detection in wastewater, suggesting ongoing community burden. Wide confidence intervals, lack of quantitative data, and limited data collection after the matches limit our conclusions. This study demonstrates the use of qualitative, multiplexed PCR assays for WBS during a mass gathering sporting event.

**Importance:** Mass gatherings can have numerous public health implications, including facilitating the spread of infectious diseases. Surveillance, both with clinical tracking of cases and through alternative methods, such as wastewater-based surveillance, can inform bio-preparedness efforts. Our study demonstrates the use of qualitative PCR assays to capture population-level pathogen information surrounding the 2026 FIFA World Cup matches held in Atlanta, GA.

## Introduction

The Fédération Internationale de Football Association (FIFA) World Cup 2026™ (FWC’26) was the largest World Cup to date, spread across the United States of America, Canada, and Mexico. Sixteen cities held matches between June 11^th^ and July 19^th^, 2026, with a total of 48 participating national teams (1). Events of this scale constitute mass gatherings (MGs), defined by the World Health Organization (WHO) as events attended by a significant number of people that have the potential to strain a host community’s planning and response resources (2). Potential consequences of MGs may include crowd crush, fire, violence, and the spread of infectious diseases (3). Among infectious diseases, respiratory and gastrointestinal (GI) illnesses are the most commonly reported at MGs, often associated with crowded conditions and poor sanitation and hygiene (4,5). Moreover, infectious diseases introduced or amplified by MGs may have global impacts as attendees disperse, especially when the host site is a major airline hub (6). Given these characteristics, MGs require multidisciplinary risk assessment and robust disease surveillance (7).

Clinical surveillance remains a cornerstone of assessing infectious disease burden during MGs (8). However, clinical surveillance also has limitations, including missing asymptomatic cases and variable access to diagnostic testing and care (9). Complementary surveillance modalities are therefore essential to fully capture the true burden of circulating pathogens and inform bio-preparedness efforts. One such method is wastewater-based surveillance, which can capture excreted pathogens regardless of patient symptoms or reporting from healthcare institutions (10). Due to these benefits, wastewater-based epidemiology gained increased attention during the COVID-19 pandemic, with many research efforts focusing on the detection of SARS-CoV-2 in wastewater (11). For example, wastewater-based surveillance was used in the Tokyo 2020 Olympics and Paralympics and the FIFA World Cup Qatar 2022 to monitor the presence of SARS-CoV-2, as well as human enterovirus and polio in the latter (12,13).

In this study, we utilize wastewater-based surveillance to examine the influence of the FWC’26 on the range of infectious pathogens in the greater Metro Atlanta area. Analysis includes a temporal comparison of pathogen detection before and during the eight FWC’26 matches held in Atlanta, GA between June 15 and July 15, 2026, using the BIOFIRE^®^ WATCHFIRE™ Respiratory and Gastrointestinal PCR Pathogen Panels.

## Methods

### 1. Wastewater sampling

The city of Atlanta hosted eight matches between June 15^th^ and July 15^th^, including one semifinal, at the Mercedes-Benz Stadium (renamed the Atlanta Stadium for the duration of the tournament). Total match attendance during this time exceeded 540,000, with an average of 68,000 attendees per match (14). Samples were collected at three water reclamation facilities in the Atlanta metro area between April 1^st^ and July 29^th^ of 2026 (n= 102, across three facilities and 34 collection days). Facilities were chosen based on their service areas and proximity to Mercedes-Benz Stadium. They are owned and operated by the City of Atlanta and have been de-identified in this study to preserve facility anonymity (Table 1).

**Table 1.** Characteristics of the three sampled water reclamation facilities.

| Treatment facility | Treatment capacity (MGD)<sup>1</sup> | Population served |
|---|---|---|
| Facility A | 122 | 600,000 |
| Facility B | 54 | 150,000 |
| Facility C | 44 | 90,000 |

<sup>1</sup>MGD = million gallons per day

**Table 2.** Target analytes for WATCHFIRE™ Gastrointestinal PCR Panel.

| **Bacterial** | **Viral** | **Parasitic** |
|---|---|---|
| *Campylobacter* | Adenovirus F 40/41 | *Cryptosporidium* |
| *Clostridium difficile toxin A/B* | Astrovirus | *Cyclospora cayetanensis* |
| *Plesiomonas shigelloides* | Norovirus GI/GII | *Entamoeba histolytica* |
| *Salmonella* | Rotavirus A | *Giardia lamblia* |
| *Vibrio* | Sapovirus |  |
| *Vibrio cholerae* |  |  |
| *Yersinia enterocolitica* |  |  |
| Enteroaggregative *E. coli* (EAEC) |  |  |
| Enteropathogenic *E. coli* (EPEC) |  |  |
| Enterotoxigenic *E. coli* (ETEC) lt/st |  |  |
| Shiga-like toxin-producing *E. coli* (STEC) stx1/stx2 |  |  |
| *E. coli* O157 |  |  |
| *Shigella*/Enteroinvasive *E. coli* (EIEC) |  |  |

**Table 3.** Target analytes for WATCHFIRE™ Respiratory PCR Panel.

| **Bacterial** | **Viral** |
|---|---|
| *Bordetella parapertussis* (IS *1001*) | Adenovirus |
| *Bordetella pertussis* (*ptxP*) | Coronavirus 229E (HCoV-229E) |
| *Chlamydia pneumoniae* | Coronavirus HKU1 (HCoV-HKU1) |
| *Mycoplasma pneumoniae* | Coronavirus NL63 (HCoV-NL63) |
|  | Coronavirus OC43 (HCoV-OC43) |
|  | Severe Acute Respiratory Syndrome Coronavirus 2 (SARS-CoV-2) |
|  | Human Metapneumovirus (hMPV) |
|  | Human Rhinovirus/Enterovirus |
|  | Influenza A |
|  | Influenza B |
|  | Parainfluenza Virus 1 |
|  | Parainfluenza Virus 2 |
|  | Parainfluenza Virus 3 |
|  | Parainfluenza Virus 4 |
|  | Respiratory Syncytial Virus |

Twice weekly during the collection period, 24-hour composite compliance samples from each of the three water reclamation facilities were brought to the lab at one of the water reclamation facilities. 250ml of primary influent (PI) wastewater was collected per facility per collection day (6 samples total per week). 1ml of this was aliquoted for raw wastewater WATCHFIRE™ testing. Samples were refrigerated and transported to Emory University Rollins School of Public Health for processing within one hour of aliquoting. Of note, we attempted to use microbe-concentration to compare raw and concentrated samples, but due to a human error in the concentration processing step, were unable to do so.

### 2. BIOFIRE^®^ WATCHFIRE™ testing

Wastewater samples from each collection date throughout the collection period were tested on the BIOFIRE^®^ FILMARRAY^®^ TORCH system, integrated with FIREWORKS™ software using the WATCHFIRE™ Respiratory and Gastrointestinal PCR Panels. The BIOFIRE^®^ WATCHFIRE™ Gastrointestinal and Respiratory Panels (made commercially available in April 2026) enable streamlined wastewater-based polymerase chain reaction (PCR) testing (12). These panels provide a qualitative, multiplexed, nested nucleic acid-based test identifying the following targets:

Briefly, 300 uL samples of raw wastewater were injected into sample buffer ampoules and then injected into the WATCHFIRE™ testing pouches. Hydration solution was injected into the testing pouch to hydrate the lyophilized reagents within the pouch. RNA Process Control and PCR2 Controls were included with each testing pouch. If internal pouch controls failed, pouches were considered invalid and were excluded from analysis. PCR testing yielded qualitative (detected/not detected, or N/A) results for each panel target per sample.

All panel targets were reported as either detected or not detected. N/A results only applied to *Escherichia coli* (*E. coli*) O157 and enteropathogenic *E. coli* (EPEC). The presence of these bacteria was reported in the context of a positive Shiga toxin-producing *E. coli* (STEC) result. *E. coli* O157 was only reported to be detected if STEC was detected. If STEC was not detected, *E. coli* O157 was reported as N/A. For EPEC, the assay was unable to determine the difference between the presence of STEC and co-contamination with EPEC and STEC. Therefore, if STEC was detected in the sample, EPEC would be reported as N/A.

### 4. Data analysis

RStudio (Version 2026.08.0+187) was utilized for all statistical analysis and graph development (16). Temporal patterns were analyzed, comparing wastewater pathogen profiles before and during the FWC’26 across the three reclamation facilities. All N/A results (see above) were treated as not detected for the statistical analysis. *V. cholerae* was excluded from all analyses and results due to inconsistencies in testing.

## Results

Wastewater was collected twice weekly from three wastewater reclamation facilities across the Atlanta area between April 1 and July 29, 2026. Overall, detection of pathogens was more prevalent on the gastrointestinal panel compared to the respiratory panel (Figure 1, 2). Heat maps for individual facilities are included in the supplemental materials.

**Figure 1.**
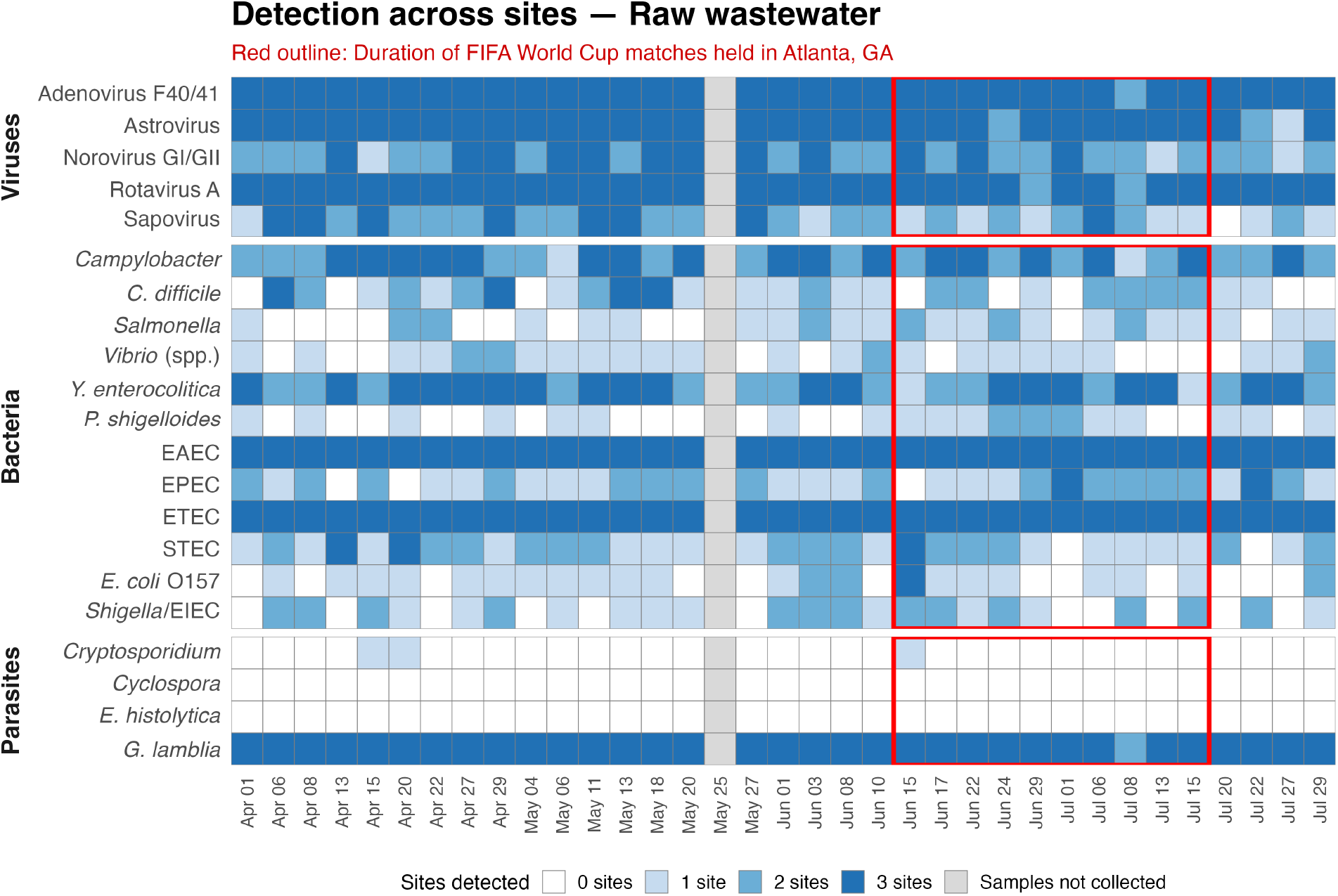
Detection of pathogens on WATCHFIRE™ Gastrointestinal PCR Panel for raw wastewater samples across all three facilities. Data was not collected on May 25, 2026. The red box delineates the days World Cup matches were being held in Atlanta, GA.

**Figure 2.**
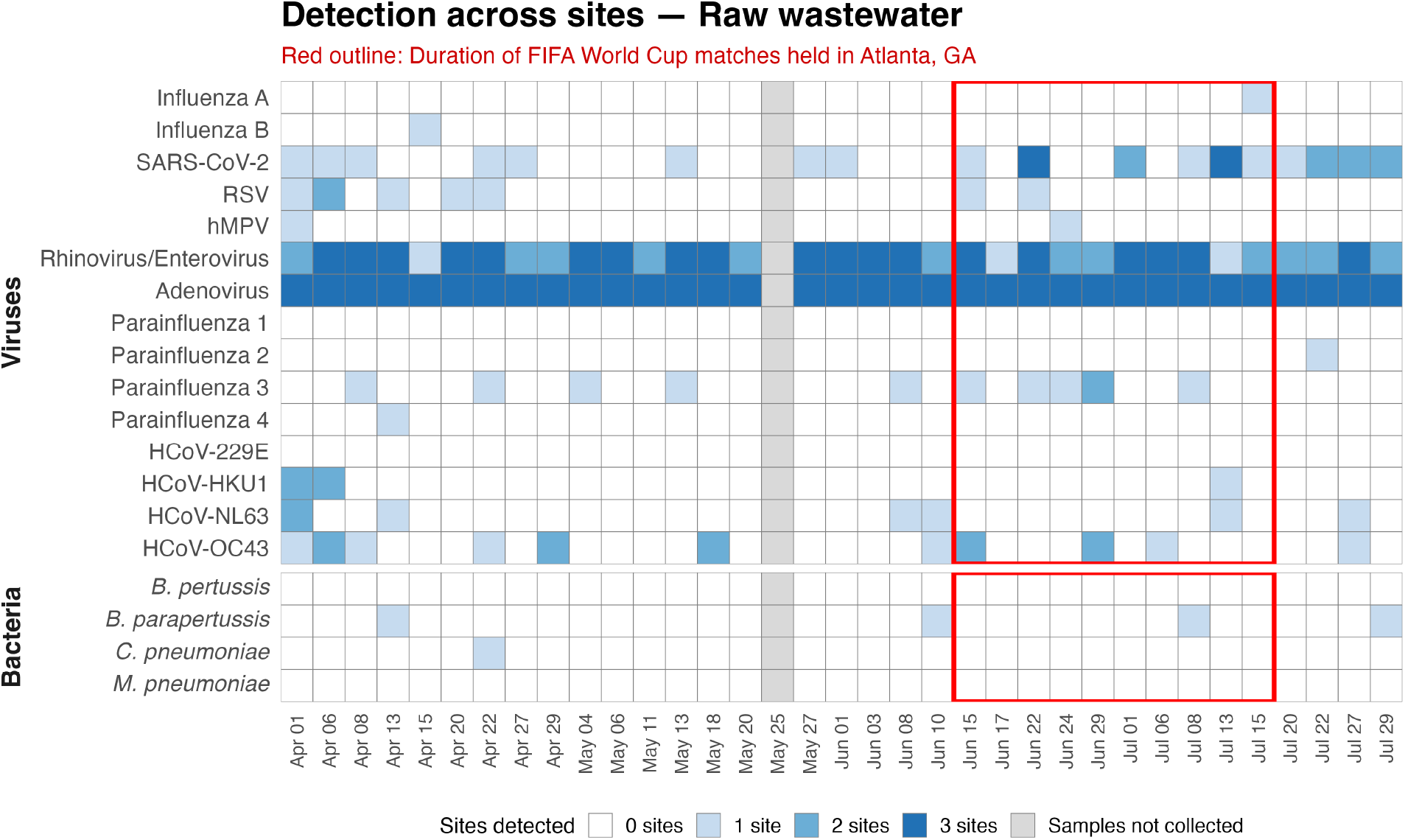
Detection of pathogens on WATCHFIRE™ Respiratory PCR Panel for raw wastewater samples across all three facilities. Data was not collected on May 25, 2026. The red box delineates the days World Cup matches were being held in Atlanta, GA.

The most commonly detected pathogens across all three reclamation facilities included EAEC and ETEC on the GI panel and adenovirus and rhinovirus/enterovirus on the respiratory panel. Least commonly detected pathogens included *Cyclospora* and *E. histolytica* on the GI panel and *B. pertussis*, *M. pneumoniae*, and parainfluenza virus 1 on the respiratory panel, none of which were detected throughout the collection period (Table 4, 5).

**Table 4.** Gastrointestinal pathogen detection frequencies on WATCHFIRE™ PCR panels before (n = 60) and during (n= 30) the FWC’ 26 pooled across three water reclamation facilities.

| **Target** |  | **Before the FWC' 26**<br>N (%) | **During the FWC' 26**<br>N (%) |
|---|---|---|---|
| Viruses | Adenovirus F40/41 | 60 (100) | 29 (97) |
|  | Astrovirus | 60 (100) | 29 (97) |
|  | Norovirus GI/GII | 48 (80) | 22 (73) |
|  | Rotavirus A | 60 (100) | 28 (93) |
|  | Sapovirus | 45 (75) | 16 (53) |
| Bacteria | *Campylobacter* | 50 (83) | 24 (80) |
|  | *C. difficile* | 30 (50) | 13 (43) |
|  | *Salmonella* | 14 (23) | 12 (40) |
|  | *Vibrio* | 18 (30) | 6 (20) |
|  | *Y. enterocolitica* | 52 (87) | 23 (77) |
|  | *P. shigelloides* | 8 (13) | 11 (37) |
|  | EAEC | 60 (100) | 30 (100) |
|  | EPEC | 27 (45) | 16 (53) |
|  | ETEC | 60 (100) | 30 (100) |
|  | STEC | 33 (55) | 14 (47) |
|  | *E. coli* O157 | 16 (27) | 9 (30) |
|  | *Shigella*/EIEC | 21 (35) | 12 (40) |
| Parasites | *Cryptosporidium* | 2 (3) | 1 (3) |
|  | *Cyclospora* | 0 (0) | 0 (0) |
|  | *E. histolytica* | 0 (0) | 0 (0) |
|  | *G. lamblia* | 60 (100) | 29 (97) |

**Table 5.** Respiratory pathogen detection frequencies on WATCHFIRE™ PCR panels before (n = 60) and during (n = 30) the FWC’ 26 pooled across three water reclamation facilities.

| Target |  | **Before the FWC' 26**<br>N (%) | **During the FWC' 26**<br>N (%) |
|---|---|---|---|
| Viruses | Influenza A | 0 (0) | 1 (3) |
|  | Influenza B | 1 (2) | 0 (0) |
|  | SARS-CoV-2 | 8 (13) | 11 (37) |
|  | RSV | 6 (10) | 2 (7) |
|  | hMPV | 1 (2) | 1 (3) |
|  | Rhinovirus/Enterovirus | 52 (87) | 23 (77) |
|  | Adenovirus | 60 (100) | 30 (100) |
|  | Parainfluenza 1 | 0 (0) | 0 (0) |
|  | Parainfluenza 2 | 0 (0) | 0 (0) |
|  | Parainfluenza 3 | 5 (8) | 6 (20) |
|  | Parainfluenza 4 | 1 (2) | 0 (0) |
|  | HCoV-229E | 0 (0) | 0 (0) |
|  | HCoV-HKU1 | 4 (7) | 1 (3) |
|  | HCoV-NL63 | 5 (8) | 1 (3) |
|  | HCoV-OC43 | 10 (17) | 5 (17) |
| Bacteria | *B. pertussis* | 0 (0) | 0 (0) |
|  | *B. parapertussis* | 2 (3) | 1 (3) |
|  | *C. pneumoniae* | 1 (2) | 0 (0) |
|  | *M. pneumoniae* | 0 (0) | 0 (0) |

Primary data analysis compared the frequency of pathogen target detections in the during-tournament period (June 15 to July 15) to the before-tournament period (April 1 to June 14). The former was defined by the dates matches were held in Atlanta, GA. The before-tournament period included 20 sample collection days per facility (n=60), excluding May 25 when data was not collected due to the Memorial Day holiday. The during-tournament period included 10 sample collection days per facility (n=30).

A modified Poisson model (Poisson-with-log-link) was utilized to calculate risk ratios (RRs) directly from a binary outcome (17). To account for differences between facilities, collection facility was included as a fixed covariate. To account for variability between collection days, collection day was included as a random effect. The during-tournament period was compared to the before-tournament period where RR >1 indicated increased detection of pathogen targets during the FWC’26. We also used the Bonferroni correction to calculate adjusted p-values to account for multiple testing.

Only *P. shigelloides* (RR = 2.75, 95% CI = 1.11-6.84, p = 0.03) showed significant change during the FWC’26 for GI pathogens (Table 6). However, this difference was not statistically significant after Bonferroni correction (p-adjusted = 0.531). Four pathogens could not be modeled because of either detection in every sample or in no sample, resulting in zero variance in outcome during the two time periods. All other GI pathogens showed no statistically significant difference between the two time periods.

**Table 6.** Comparison of during-tournament GI pathogen detection to before-tournament detection.

| **Target** |  | **RR (95% CI)** | **Unadjusted p-value** | **Note** |
|---|---|---|---|---|
| Viruses | Adenovirus F40/41 | 0.97 (0.62-1.51) | 0.881 | - |
|  | Astrovirus | 0.97 (0.62-1.51) | 0.881 | - |
|  | Norovirus GI/GII | 0.92 (0.55-1.52) | 0.735 | - |
|  | Rotavirus A | 0.93 (0.60-1.46) | 0.763 | - |
|  | Sapovirus | 0.71 (0.40-1.26) | 0.242 | - |
| Bacteria | *Campylobacter* | 0.96 (0.59-1.56) | 0.869 | - |
|  | *C. difficile* | 0.87 (0.45-1.66) | 0.667 | - |
|  | *Salmonella* | 1.71 (0.79-3.71) | 0.171 | - |
|  | *Vibrio* | 0.67 (0.26-1.68) | 0.390 | - |
|  | *Y. enterocolitica* | 0.88 (0.54-1.45) | 0.624 | - |
|  | *P. shigelloides* | 2.75 (1.11-6.84) | 0.030 | - |
|  | EAEC | - | - | Zero variance in outcome<sup>2</sup> |
|  | EPEC | 1.19 (0.64-2.20) | 0.590 | - |
|  | ETEC | - | - | Zero variance in outcome |
|  | STEC | 0.85 (0.45-1.59) | 0.607 | - |
|  | *E. coli* O157 | 1.13 (0.50-2.55) | 0.777 | - |
|  | *Shigella*/EIEC | 1.14 (0.56-2.32) | 0.712 | - |
| Parasites | *Cryptosporidium* | 1.00 (0.09-11.03) | 1.000 | - |
|  | *Cyclospora* | - | - | Zero variance in outcome |
|  | *E. histolytica* | - | - | Zero variance in outcome |
|  | *G. lamblia* | 0.97 (0.62-1.51) | 0.881 | - |

### 2 Pathogen detected in samples from every collection day or no collection day across both time periods

For the respiratory pathogens, only SARS-CoV-2 had an increase in the during-tournament period (RR = 2.75, 95% CI = 1.11-6.84; p = 0.03) (Table 7). However, this difference was also not significant after Bonferroni correction (p-adjusted = 0.383). Meanwhile, six pathogens had zero variance in outcome, and risk ratios for four pathogens could not be estimated due to low detection and unbounded confidence intervals.

**Table 7.** Comparison of during-tournament respiratory pathogen detection to before-tournament detection.

| **Target** |  | **RR (95% CI)** | **Unadjusted p-value** | **Note** |
|---|---|---|---|---|
| Viruses | Influenza A | - | - | Not estimable |
|  | Influenza B | - | - | Not estimable |
|  | SARS-CoV-2 | 2.75 (1.11-6.84) | 0.030 | - |
|  | RSV | 0.67 (0.13-3.30) | 0.620 | - |
|  | hMPV | 2.00 (0.13-31.98) | 0.624 | - |
|  | Rhinovirus/Enterovirus | 0.88 (0.54-1.45) | 0.624 | - |
|  | Adenovirus | - | - | Zero variance in outcome |
|  | Parainfluenza 1 | - | - | Zero variance in outcome |
|  | Parainfluenza 2 | - | - | Zero variance in outcome |
|  | Parainfluenza 3 | 2.40 (0.73-7.86) | 0.148 | - |
|  | Parainfluenza 4 | - | - | Not estimable |
|  | HCoV-229E | - | - | Zero variance in outcome |
|  | HCoV-HKU1 | 0.83 (0.01-137.63) | 0.944 | - |
|  | HCoV-NL63 | 0.41 (0.04-4.40) | 0.464 | - |
|  | HCoV-OC43 | 0.99 (0.29-3.42) | 0.988 | - |
| Bacteria | *B. pertussis* | - | - | Zero variance in outcome |
|  | *B. parapertussis* | 1.00 (0.09-11.03) | 1 | - |
|  | *C. pneumoniae* | - | - | Not estimable |
|  | *M. pneumoniae* | - | - | Zero variance in outcome |

A sensitivity analysis was performed, extending the during-tournament period until July 29^th^. Similar to the primary analysis, this analysis also showed an increase in *P. shigelloides* (RR = 2.50, 95% CI = 1.05-5.96; p = 0.04, p-adjusted = 0.696) which was not statistically significant after Bonferroni correction. For the respiratory panel, SARS-CoV-2 still had a statistically significant increase in the during-tournament period compared to before the tournament with a higher RR than in the primary analysis (RR = 3.21, 95% CI = 1.40-7.39; p = 0.01, p-adjusted = 0.048). There were no other statistically significant differences between defining the during-tournament period as June 15 to July 15 compared to June 15 to July 29.

## Discussion

In this study, we characterized the pathogen composition of three Atlanta water reclamation facilities before and during the FWC’26 using the BIOFIRE^®^ WATCHFIRE™ Respiratory and Gastrointestinal PCR Pathogen Panels. Overall, pathogen composition of wastewater across three Atlanta reclamation facilities did not substantially change before and during the FWC’26.

Though there were observed increases in SARS-CoV-2 and *P. shigelloides* during the FWC’26 compared to before, these were not statistically significant.

Increased detection of SARS-CoV-2 in our sensitivity analysis, which was statistically significant, could be explained by an increase in regional and national detection during the latter half of the summer as documented by the CDC (18,19). It is difficult to state whether the increase in SARS-CoV-2 observed can be attributed to the FWC’26 given overall increased detection during the same period, which suggests an ongoing community burden as the more likely explanation for our findings.

Several characteristics of the FWC’26 likely mitigated the risk of communicable disease spread, including scattered accommodations across the city, short interval matches after which attendees disperse, and established city infrastructure (20). These factors may have contributed to the overall lack of change in disease incidence observed in our results. However, our wide confidence intervals also suggest a moderate degree of uncertainty, limiting the conclusions we can draw from our data. Of note, there were no reports of gastrointestinal or respiratory illness outbreaks by the Georgia Department of Public Health during the FWC’26 either (21).

Overall, limitations of this study included limited sample size with only three months of sample collection and often low frequency of pathogen detection, particularly on the respiratory panel. Multiple pathogens on both the GI and respiratory panels were unable to be modeled due to limited detection. We also had less availability of data after the FWC’26 ended compared to data before the event. Moreover, without quantitative pathogen data, results are limited to comparison of binary detection before and during the World Cup without being able to determine if the pathogen burden had increased. Lastly, using a PCR panel limits pathogen detection of panel targets, missing other potential targets that may be captured by metagenomic sequencing. The WATCHFIRE™ PCR panels are also newer assays that require continued testing, as research has shown false-positive rates for certain GI viruses when compared to RT-qPCR and conventional RT-PCR methods (22).

In summary, this study utilized PCR assays to conduct wastewater-based surveillance of GI and respiratory pathogens during a summer sports mass gathering event. Pathogen composition remained largely unchanged during the event, and the few changes that were observed were not statistically significant or had competing, alternative explanations. However, PCR-based wastewater epidemiology remains a useful tool in quickly characterizing community infection prevalence of a wide array of pathogens, especially with expanded panels that enable detection of multiple pathogens simultaneously. Having additional quantitative data and metagenomic sequencing can be further used to comprehensively assess pathogen burden in a community, especially when there is increased risk of communicable disease spread such as mass gathering events.

## Acknowledgements

CSK is a medical advisor for Ferring/Rebiotix, scientific ad board for Vedanta Biosciences, Seres Therapeutics.

This project was supported by the National Center for Advancing Translational Sciences of the National Institutes of Health under Award Number UL1TR002378. The content is solely the responsibility of the authors and does not necessarily represent the official views of the National Institutes of Health.

## Supplemental Materials

**Supplemental Figure 1.**
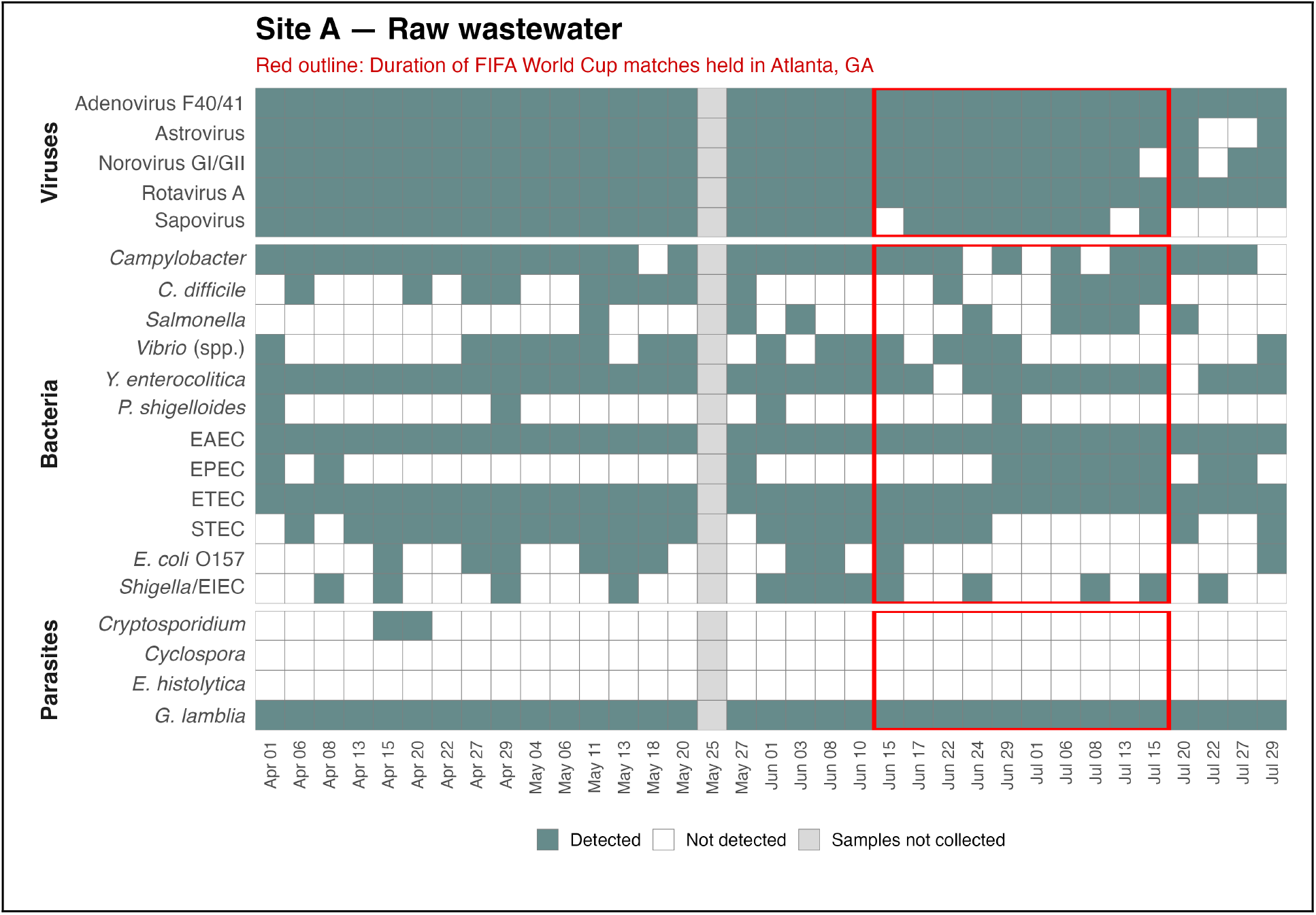
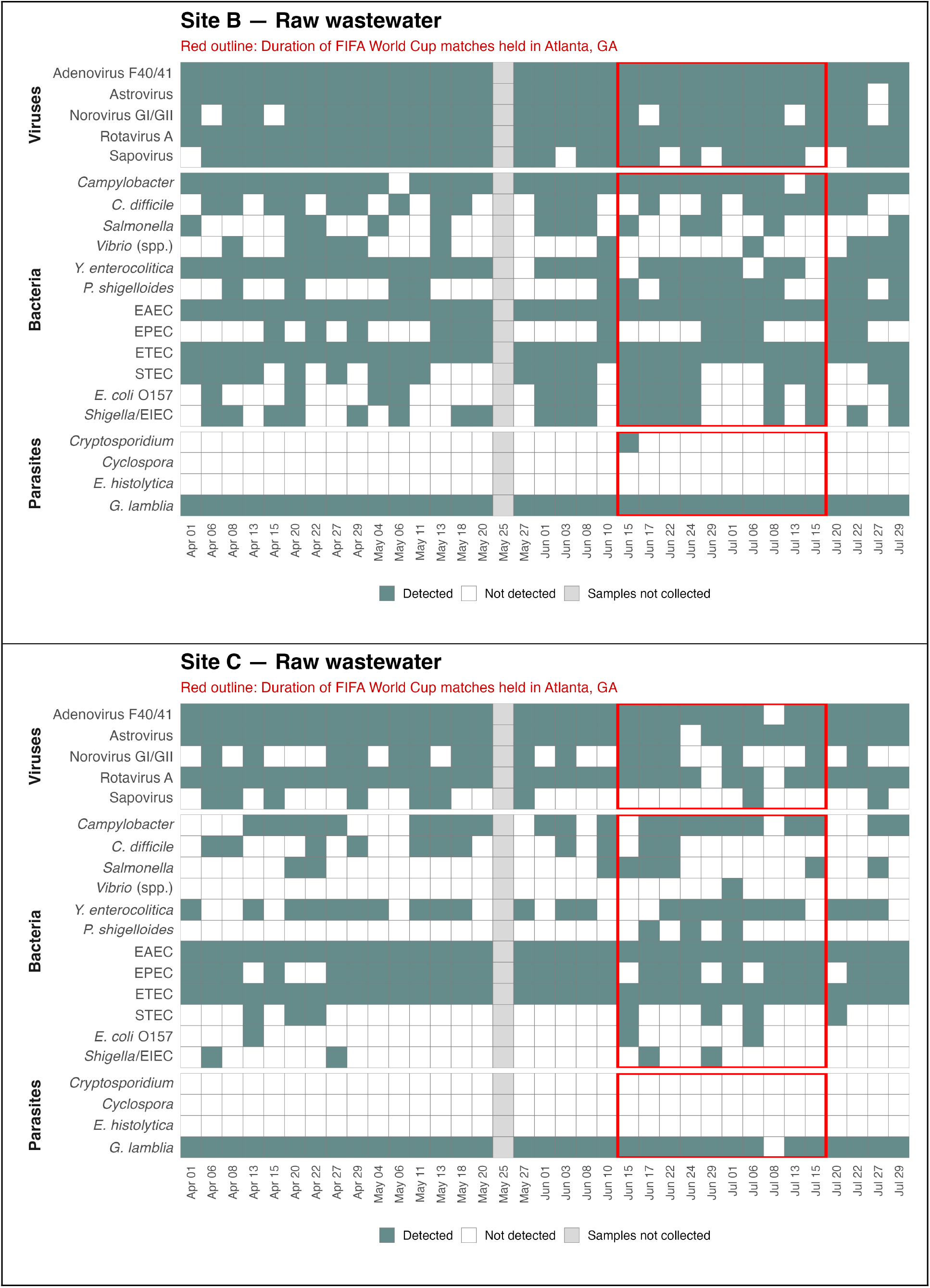
Detection of pathogens on WATCHFIRE™ Gastrointestinal PCR Panel for raw wastewater samples per facility.

**Supplemental Figure 2.**
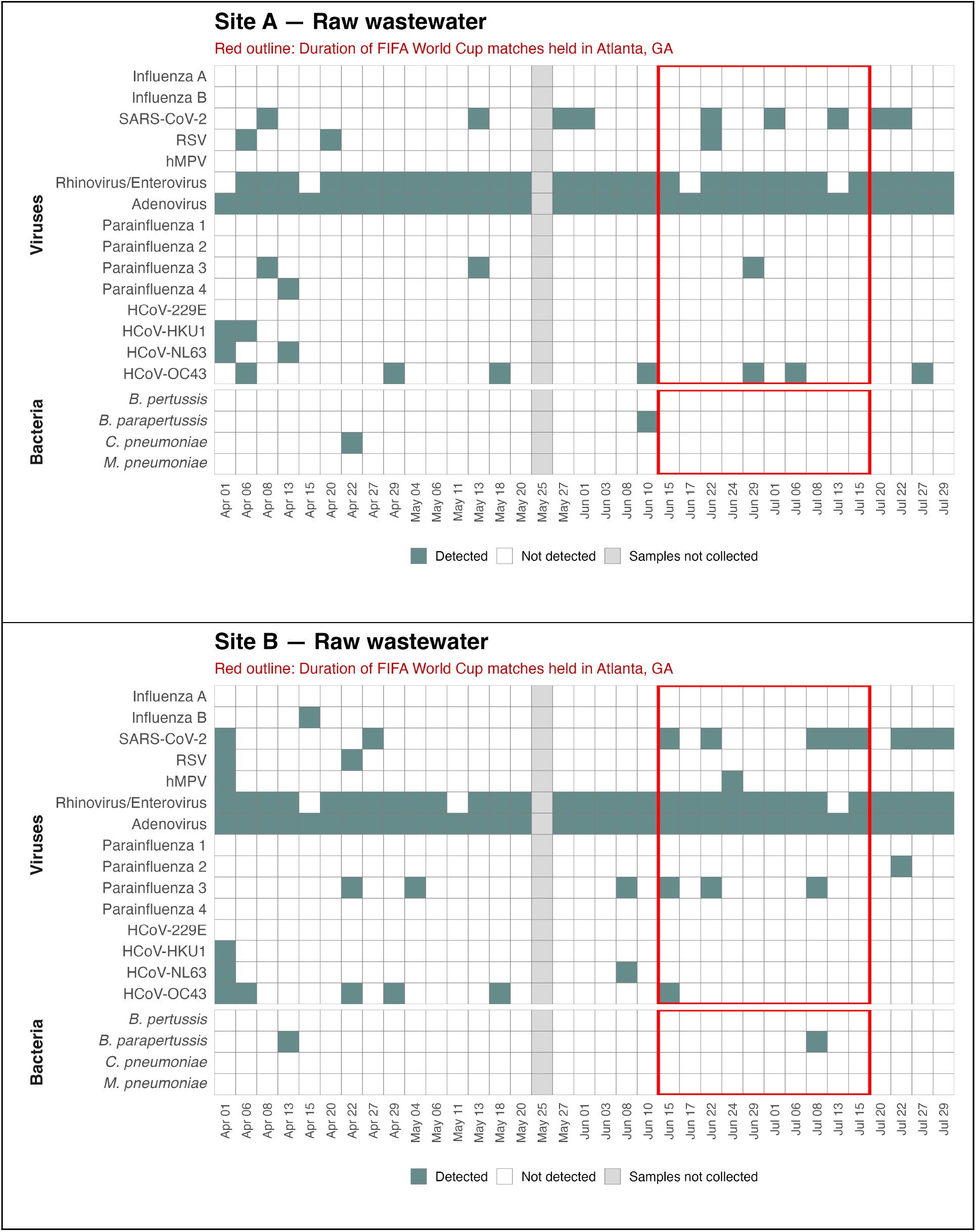
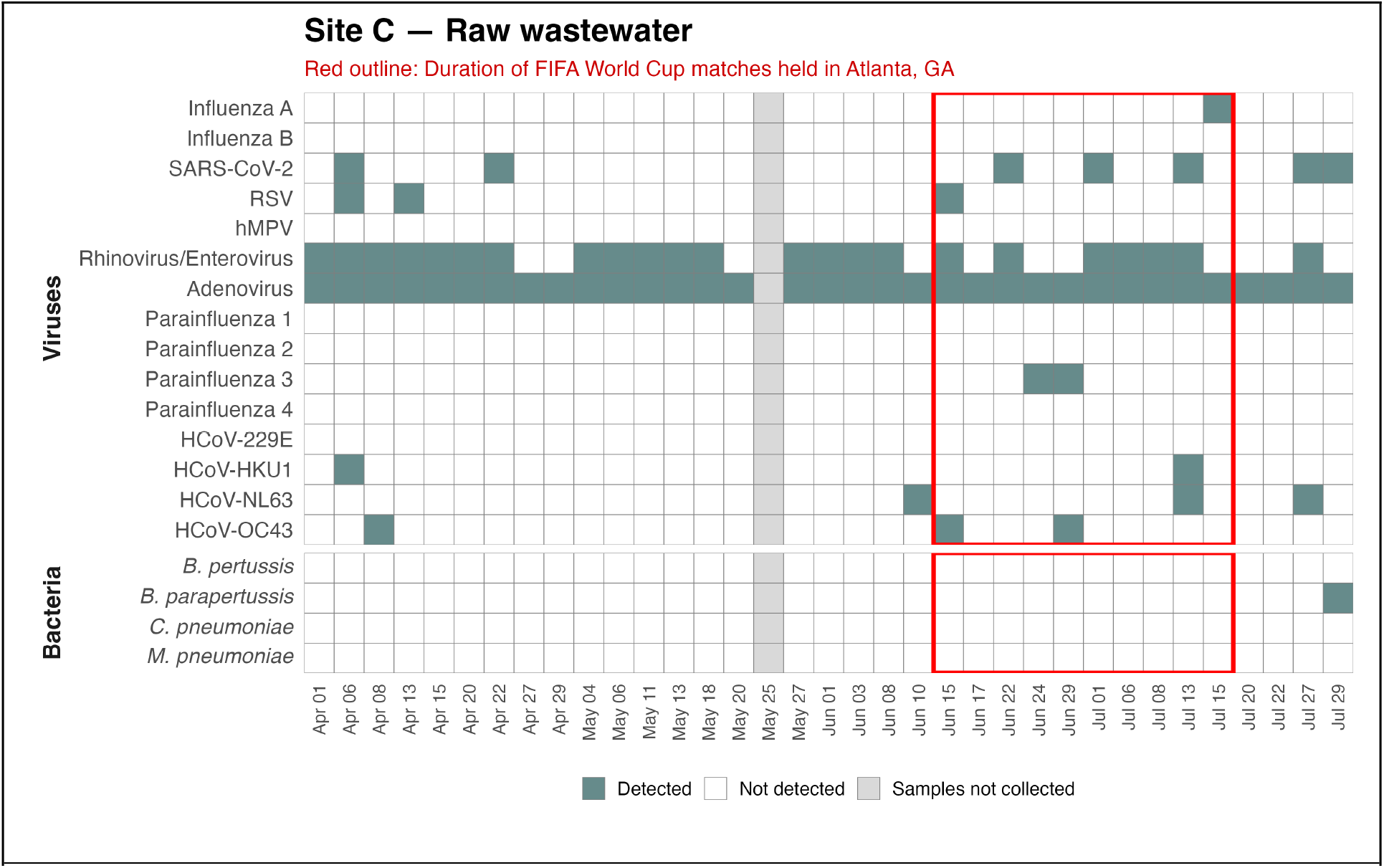
Detection of pathogens on WATCHFIRE™ Respiratory PCR Panel for raw wastewater samples per facility.

### Data Availability

The raw data supporting these findings are not publicly available due to confidentiality agreements.

